# “It is guiding us to protect ourselves”: a qualitative investigation into why young people engage with a mass-media HIV education campaign

**DOI:** 10.1101/2022.04.05.22272126

**Authors:** Venetia Baker, Sarah Mulwa, Sophie Sarrassat, David Khanyile, Cherie Cawood, Simon Cousens, Isolde Birdthistle

## Abstract

This study explores how and why young people engage with MTV Shuga, a popular mass media edutainment campaign, to understand what makes effective HIV education. Young MTV Shuga viewers from the Eastern Cape, South Africa and their parents participated in remote individual interviews and focus groups in 2020. Qualitative data were transcribed and analysed using a thematic iterative approach. Young participants engaged with MTV Shuga for relatable, tolerant, and complex stories about young people navigating HIV and relationships. These stories, which made viewers aware of sexual health services, inspired young people to reflect on how they would approach sexual health scenarios. MTV Shuga initiated conversations among peers, partners and some families about HIV that made them feel supported and equipped to tackle problems in their own lives. Storytelling can make HIV education engaging and relatable as it guides young people through sexual health situations. Storytelling can empower young people to make their own informed decisions while allowing space for uncertainty and diverse opinions about HIV and sexual health. Media-based storytelling can disseminate knowledge into homes and start conversations about HIV in the broader community. Increasing digitally-unconnected youths’ access to media-based interventions is essential to making HIV education more equitable.

## Introduction

South Africa, the country with the largest HIV epidemic, has documented encouraging declines in HIV incidence in recent years. This is largely attributed to major gains in treatment access and resulting viral suppression among people living with HIV, along with voluntary male medical circumcision (I. Birdthistle, Kwaro, et al. 2021). Yet new cases remain high in young people aged 15 to 24 years, at an estimated 88,400 new infections in 2017 or 1.0% per year, with a substantial difference between males and females, 0.49% and 1.51% respectively (*South African National HIV Prevalence, Incidence, Behaviour and Communication Survey*, 2017). Self-reported HIV testing is about 10% lower in young males and females aged 15-24 years old (39% and 53% respectively) compared to 25 to 49 years old males and females (48% and 62% respectively)(*South Africa Demographic and Health Survey* 2016:). Despite investments, comprehensive knowledge of HIV among young people remained low over the past decade(De Wet, Akinyemi, and Odimegwu 2019). A 2017 Human Sciences Research Council survey reports that just over a third of young people (36.1 %) aged 15-24 years displayed accurate knowledge about the sexual transmission of HIV and rejected major misconceptions about HIV.

One behavioural health education campaign that has shown to increase HIV knowledge and improve other sexual health outcomes is MTV Shuga. *MTV Shuga* is a mass media campaign created by the Staying Alive Foundation, centred around a popular TV drama that aims to educate young people about sexual health, particularly HIV prevention and testing, using relatable storylines. Over the past decade, millions of young people have been motivated to seek out, watch, and engage with this edutainment campaign. Series have been located across sub-Saharan Africa in Kenya, Nigeria, Cote d’Ivoire and South Africa. *MTV Shuga* has reached an estimated 719 million households through 179 terrestrial channels and another 42 million viewers through rights-free streaming on the internet. As of November 2021, the premiering episode of MTV Shuga Down South, the South African series, had over 700 thousand viewers on YouTube with 5 thousand likes and 500 comments on that platform. Several studies have shown that exposure to *MTV Shuga* is associated with greater awareness and motivation to adopt HIV prevention practices and improvements in other sexual health outcomes, including greater likelihood to use condoms at last sex, lower incidence of teenage pregnancy and less tolerance for gender-based violence (Banerjee, Ferrara, and Orozco-Olvera 2019; Isolde Birdthistle, Mulwa, et al. 2021; Shahmanesh et al. 2019). A trial in Nigeria found increases in HIV and STI testing among study participants in communities randomly selected to receive group viewings of *MTV Shuga* episodes (Banerjee, Ferrara, and Orozco-Olvera 2019). Among a cohort of adolescent girls and young women in rural KwaZulu-Natal, South Africa exposure to *MTV Shuga* was relatively low but associated with a range of sexual and reproductive health benefits (Shahmanesh et al. 2019). Most recently, a mixed-methods evaluation in the Eastern Cape of South Africa found that knowledge of HIV status and awareness of HIV self-screening and PrEP was higher among those exposed to MTV Shuga compared to those with no exposure to the campaign (Isolde Birdthistle, Mulwa, et al. 2021).

Social and behaviour change communication programmes that use radio, television, and other forms of entertainment media have been proposed as an innovative solution to improve HIV knowledge and promote healthy sexual practices (*South African National HIV Prevalence, Incidence, Behaviour and Communication Survey*, 2017). Though research has evaluated the effectiveness of mass media on HIV related outcomes (Vidanapathirana et al. 2005; Bertrand et al. 2006; Stead et al. 2019), less is known about why people engage with mass-media edutainment campaigns and how it influences them. MTV Shuga is a compelling intervention to examine because millions of young people have actively sought out the series. This analysis explores how and why young people engage with MTV Shuga to understand what makes it an effective and engaging sex education programme.

## Methods

### Study setting

The South African Department of Health recommended that this research be conducted in the Eastern Cape of South Africa where the prevalence of HIV has been estimated at 15% (*South African National HIV Prevalence, Incidence, Behaviour and Communication Survey*, 2017). There is relatively low HIV testing uptake among young people (15-24 years old). According to the 2016 Demographic and Health Survey (DHS), 48% (95%CI 33, 64) and 56% (95%CI 49, 64) of young males and females in urban settings of Eastern Cape province had tested for HIV and received their result in the past 12 months (*South Africa Demographic and Health Survey* 2016:). Lower proportions were found in the males and females aged 15 to 19 years old: 35% (95%CI 20, 52) and 42% (95%CI 32, 54) respectively (*South Africa Demographic and Health Survey* 2016:). The city of Mthatha was chosen as a study setting because of the opportunity to engage with *MTV Shuga*. According to the 2016 DHS, 79% (95%CI 73, 84) of households in urban settings of Eastern Cape province owned a TV and 76% (95%CI 62, 86) and 67% (95%CI 52, 79) of young males and females in urban settings of the Eastern Cape province watched TV in the past week respectively (*South Africa Demographic and Health Survey* 2016:). MTV Shuga DS2, was available on SABC (a national TV channel), local radio stations and YouTube. The MTV Shuga campaign ran peer-education and community events and distributed a graphic novel through schools in Mthatha.

### Eligibility and sampling

Participants were recruited from an online survey about *MTV Shuga* that targeted young people living in Mthatha (Isolde Birdthistle, Mulwa, et al. 2021). Those who completed the survey could opt-in to be contacted for further qualitative research. Young people from Mthatha and the surrounding areas and exposed to at least one episode of DS2 were eligible to participate. Parents were recruited through qualitative participants 15-19 years and parent WhatsApp groups from the school that had promoted the online survey. All participants signed informed consent forms by email, and for individuals under 18 years, informed assent and informed consent of parents and guardians was required.

### Study design and data collection

In October 2020, young people aged 15-24 who reported having watched *MTV Shuga*, DS2, participated in 31 individual interviews and six age-and gender-specific focus groups of 4-6 participants. 15 parents participated in individual interviews (table 1). Interviews and focus groups were conducted remotely with individual interviews using WhatsApp and the phone, and focus groups using Zoom. Bilingual data collectors under 30 years old and trained in qualitative interviewing and remote data collection conducted the interviews. Data collectors used topic guides that were developed in English, but participants could choose to be interviewed in English, isiXhosa or Zulu. Translators transcribed the interviews into English. Questions in the topic guides were designed using The Behaviour Change Wheel, a framework for characterising behaviour change interventions (Michie, van Stralen, and West 2011).

**Table 1:**
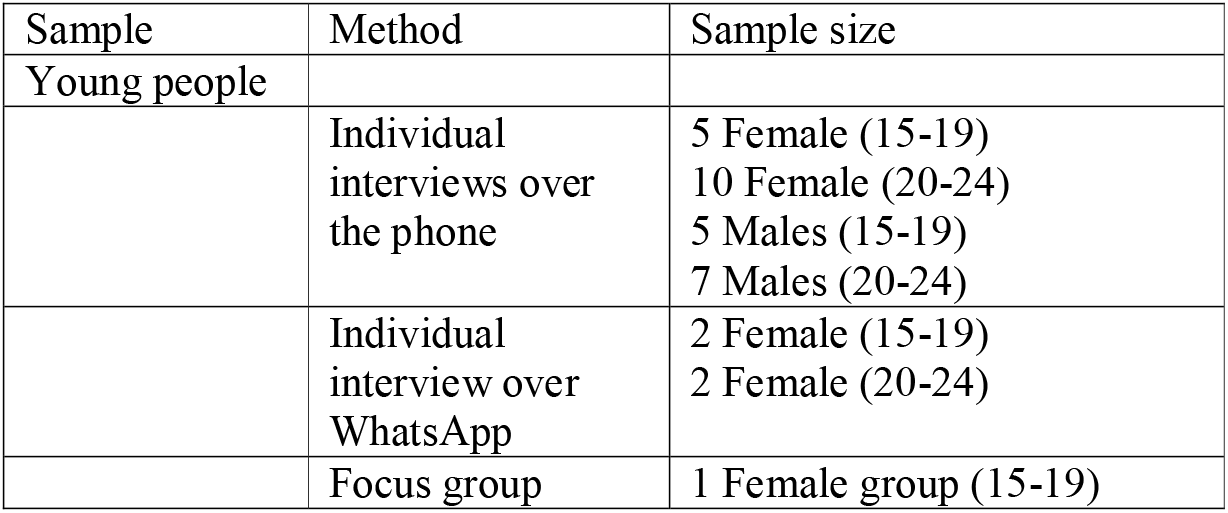
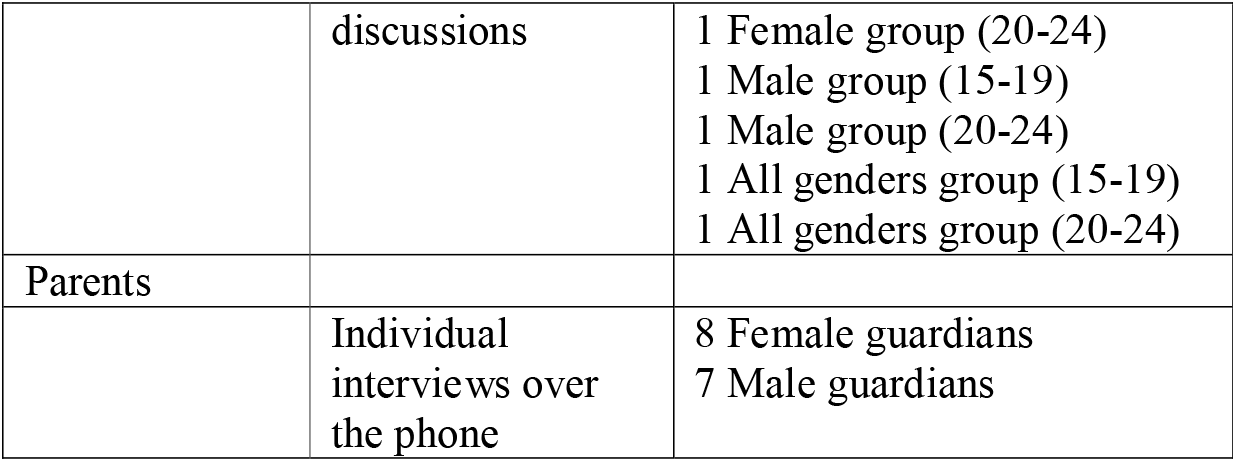
Methods design

### Analysis

Prior to the analysis, the research team considered how the data collectors’ position (Zulu and Xhosa, male and female, and under the age of 30) and remote methods might affect how participants responded to interviews and focus groups. Additionally, they reflected on how the position (White, British, female) and knowledge, attitudes and assumptions of the researcher conducting the qualitative analysis might affect the interpretation of the data.

Using a thematic iterative approach (Corbin and Strauss 2014) to analyse the data, the researcher familiarised themselves with, and coded, the transcripts. As themes and codes developed, constant comparisons were made, and divergent viewpoints that challenged emerging interpretations were identified. The researcher discussed the findings with the transcriber and data collectors to ensure the findings accurately reflected the data collected.

Quotes were extracted from the transcripts to illustrate that findings were grounded in the data. To maintain confidentiality, pseudonyms are used to attribute the quotes to young participants under 25. Pseudonyms were created using the top 30 most common registered birth forenames in the Eastern Cape province in 2017. No Pseudonyms were applied to parents who are referred to as *Parent*.

#### Findings

Young people engaged with the HIV prevention lessons in *MTV Shuga* because the series; 1) was relatable and trustworthy; 2) provided non-judgemental information about HIV and sex 3) Captured the viewers a nuanced and involved way of learning through storytelling; 4) fostered debates and shared learning; 5) Broke the ice on sexual health conversations 6) Enabled an environment for sharing sensitive sexual health information. Two significant themes emerged as barriers to engaging with the intervention; 1) young people avoided watching MTV Shuga with parents, and 2) digital inaccessibility made it difficult for some to watch and discuss.

### Reasons to engage with MTV Shuga

#### Relatable and trustworthy

Young participants felt represented by *MTV Shuga* because it accurately captured what life was like as a young person in South Africa, making them confident in *MTV Shuga* as a source of information. The series, based in Johannesburg, used current South African music, and integrated local languages and township slang.

> It’s relatable as they were South African actors. There are Xhosa characters and I’m Xhosa too. So that really kept me engaged. Just the storyline and the characters were so interesting and the language too. It’s easily relatable, it was easy to keep in tune with. - *Enzokuhle*

Additionally, participants liked that the show used young actors and was tailored to a youth audience, featuring storylines that reflected their lived experiences.

> She [the character] is my age, she’s going through the same things that I’m going through so I want to see how they make their decisions and what’s the next step. - *Alunamda*

One of the most prevalent consistent patterns that appeared in the young participant data were comments stating that *MTV Shuga* storylines reflected what was ‘happening in real life.’ Participants highlighted stories that resonated with them, such as transactional relationships, conflicts with intolerant parents, violence and masculinity, bullying, and trust in a sexual partner, as they had experienced or witnessed these situations within their own lives. When asked if they trusted the information about HIV Self-Screening and PrEP in the series, young participants regularly commented that they did because MTV Shuga, as a series, was an accurate and believable representation of their lives.

> It covers the relationships that most young people find themselves in. So, they are dating, they have social lives, and there is also an issue struggling financially… So, I think it represents or reflects what’s happening in South Africa. - *Blessing*

### Provided non-judgemental information about HIV and sex

A primary motivation for watching *MTV Shuga* was to gain more information about HIV and sexual health.

> I watched Down South because I wanted to know more about what is happening around us as youth, things we experience. And also, to have more information about HIV and AIDS. – *Princess*

Young participants were eager for information about sex because, though ‘these things are happening,’ ‘no one was talking about them.’ Participants particularly stated that parents and older people didn’t want to talk about sex and relationships with them, though participants also admitted that they avoided these conversations.

> When I was watching I learned a lot. It [*MTV Shuga*] has issues that are needed to be addressed in our community and issues that we can’t discuss with other people or older people. - *Angel*

Participants explained that *MTV Shuga* was different from other sex education because it taught them, as *Kungawo* expressed, to ‘not judge a person.’ The series raised their awareness about the situation’s others face, which made them more empathetic and tolerant. Some participants wished more people, including parents, teachers, and friends, were exposed to the show to make them more sympathetic and educated, or to feel less judged.

> I actually recommend this show to other people now so that they won’t feel judged when they’re talking about their sexuality, so that they could be more open. - *Othalive*

### Captured viewers a nuanced and involved way of learning through storytelling

The immersive experience of watching a 10-episode TV drama was also an important component that engaged young people in the show. The emotional and dramatic storylines kept participants engaged as they wondered what would happen next to the characters. Young people ‘learnt with the actors,’ as they watched them navigate scenarios, make decisions and experience the consequences of their actions.

> They [*MTV Shug*a] were not throwing or force-feeding you the information, but instead they were just explaining everything through the actors themselves, so as the actors are learning, you are learning at the same time. – *Melokuhle*
>
> I found out how each one [HIV self-screening and PrEP] works from each experience. The characters showed me the HIV testing; I saw it [HIV self-screening] from [the character] Ipeleng and her boyfriend, then I saw how PrEP works from [the character] Que’s girlfriend. - *Grace*

Participants felt empowered to form their own opinions about HIV and sexual health topics and services after absorbing the information they saw in the show. *Khayone* said he liked watching MTV Shuga because he was not forced to adhere to ‘right or wrong’ behaviours and opinions. He enjoyed the poll questions at the end of the episodes, which asking viewers their opinions about what the characters should do.

> I liked that there was an option of, ‘I’m not sure, and I don’t know.’ It wasn’t really forcing you to actually have an answer to it. And it made it interesting for you to engage [in discussions] with other people. It didn’t make you feel bad for your opinion. […] It’s not every day you are given an opportunity to say ‘I don’t know’ and I really liked that. - *Khayone*

Participants expressed that MTV Shuga storylines ‘guided them’ to make their own decisions by showing them the potential consequences of choices and the sexual health services and tools they could access. Participants hinted that this ‘guiding’ was different from the prescriptive health messages they learnt in school or from “lecturing” parents. With MTV Shuga, young people were compelled to engage in complicated storylines where characters were often not presented with an easy choice. Participants worked through the character’s situations by weighing options and engaging in debates which made them feel more prepared to face challenges in their own life. *Ncumisa* explained how she and her cousin would ‘discuss how we could have solved the particular problem that one character dealt with if it was me.’ Similarly, *Alunamda* said she watched the show so when she “comes across such [problem,] I know how to deal with it.’

### Fostered debates and shared learning

Engaging in discussions and debates seemed to be a primary component of the viewing experience and a motivator to engage with Shuga. Many young people chose to watch Shuga with a group of friends, especially among older participants living in Halls of Residence at tertiary institutions. *Likuwe* described, “everyone is quiet and concentrating so that after the show we could have like a little debate.’ In these conversations, young people often engaged in shared decision making about how they personally would react or address the situations present in the MTV Shuga storylines. One example of this was a conversation captured between *Ayanda* and *Lubanzi* during a focus group. They discussed the storyline of Que, who discovers his girlfriend Dineo has a Blesser- an older rich man who provides money and gifts you young women in exchange for sexual favours.

> I would use a different approach instead of Que’s because I’d actually sit down with Dineo, and ask her what’s going on. What would make you do what she did? Because Que took a different approach by coming on her strong, and I think that’s what made them have a misunderstanding. - *Ayanda*
>
> I agree with him [*Ayanda*]. We must try and sit down with people and try to find out the root of their problem, so we can help them, so that we do not have to sell our bodies. We must try and have another solution. - *Lubanzi*

Through these conversations, participants often explained how they were engaged on another level of learning where they learnt from their peers.

> After we have watched the show we would discuss the lifestyle and then we will compare or refer it back to us, and say how are we going to deal with this, because it’s things that are happening in our lives. Then we would say we need to take care of ourselves because most of the things that are happening in the show are relatable. So, we would teach each other about the show. - *Iminathi*

### Broke the ice on sexual health conversations

Young people explained that debates about MTV Shuga usually led to more personal conversations with friends, partners and parents about sexual health and relationships they felt they couldn’t usually discuss.

> Straight after watching the show, I had an open debate with my friends about sexual things that are happening around the youth and stuff. So, MTV Shuga helped me talk about issues that I wouldn’t even think of talking to my friends about. - *Iminathi*

One participant explained that, though it was awkward, watching MTV Shuga with her guardian allowed them to talk about HIV prevention which they wouldn’t have done if they had not watched it.

> Before, I was reluctant to discuss things with [guardian], but after watching, we would discuss the show and relate some things to real-life […] Things my mom liked talking about is knowing your status whether you’re sexually active or not, and the use of protection, and knowing your partner’s status - *Thandiswa*

Those who were open to watching MTV Shuga with their parents, like *Princess*, believed parents could ‘learn that being honest and talking about these things will make us more aware of what is going on.’ Also, *Luphawu* stated that parents could directly benefit from the series because they “do have problems like mine’ and they ‘need to know about self-screening and PrEP.’

After seeing clips of MTV Shuga in the interviews, almost all parents said they would be open to watching the show with their children. They felt it was their responsibility to educate their child about sex despite acknowledging that these conversations are often difficult and awkward.

> There is no beating around the bush about these things. You need to speak about things like this, same-sex relationships, blessers, HIV. So, I would definitely speak to them. Whether we like it or not, we have to have these conversations, and as parents, we need to play our part and be more informed and talk about these things with them. – *Parent*
>
> It is important to talk to your child about these things so when your child does engage with these things, they know the consequences. Because my mom talked to me about these things. It was also not easy for our parents to talk to us also, but it had to be done. – *Parent*

Parents were eager to have more resources and a way to break the ice to initiate these conversations. Though many stated that they thought they should discuss HIV and sexual health with their children, many had not yet done so. They felt Shuga could be a resource that could help to approach these subjects with their children.

> We are not talking about these things, black households, to be specific. […] Shuga will actually help to break the ice to help parents talk about this with their kids. So, after watching the show, you can discuss what we have learnt while we were watching. - *Parent*

Many parents learnt about HIV Self Screening and PrEP for the first time, making it easier to talk with their children about HIV prevention. As one parent explained ‘I didn’t know about PrEP and HIV self-screening. Now, I can sit down with my children and tell them about it.

### Enabled an environment for sharing sensitive sexual health information

Young participants felt safer disclosing their HIV status and other personal sexual health information to their peers after watching MTV Shuga together. Based on their discussions around MTV Shuga, they knew their friends would be supportive and understanding.

> I was happy that we got to have a discussion like that. It was long overdue for us to have because we’ve been friends for a very long time. […] It made me see that I really wouldn’t be afraid to tell my friends that I am HIV positive. […] God forbid I would find myself in such a situation where I am HIV positive, I know that I can tell my friends and they will be very supportive. - *Khayone*

Couples, in particular, explained how MTV Shuga initiated conversations around HIV testing, status disclosure, PrEP, and trust and safety within the relationship.

> You can get a head start on that thing about the HIV status of your partner and stuff because it [MTV Shuga] would be playing on TV, so you will just be doing a follow-up. – *Oyintando*
>
> [We discussed] the importance of knowing about each other’s statuses. And if you are not honest and have other girlfriends, he needs to take PrEP so that we’ll be protected. – *Alunamda*

In *Iminathi’s* relationship, watching and discussing the series had led to action as it had motivated his girlfriend and him to get tested together, sharing ‘now we both know our statuses.’

One divergent perspective was from *Luphawu* who said a conversation with friends revealed he could not trust his friends with disclosing his sexuality because of their reactions to a gay character on MTV Shuga. ‘It [the discussions] made me see that if I would tell my friends that I’m gay, they would kick me out of our friendship.’ Though the series did not make his friends more tolerant and accepting, it helped him assess that they were not safe people to disclose to.

### Barriers

#### Young people avoided watching MTV Shuga with parents

Few young participants were willing to watch the show with their parents because they feared judgment and lectures. As *Hope* explains, ‘they will think you are also doing these things that are happening on the show.’ *Linomtha* said her parent’s thought the show had a ‘bad effect on us since we are underage.’

> I would never, ever commit suicide by watching with my parents. For me, this is primarily because the show is too explicit. At home, it is because of religious reasons. Well growing up, my [guardian] prohibited us from watching soap operas. - *Gift*

Young participants were also resistant to the show becoming more parent-friendly. They worried about Shuga being ‘sugar-coated’ for a parent audience as young people deemed it too sexually explicit for their parents.

> It is too explicit for parents, but us, as youth, I feel as if we are mentally liberated enough and open-minded to understand that such ‘soapies’ [Soap operas] have now become a place for us to learn. […] So, for youth generally, it’s a very good way, but for parents and those who come from the stone age- No. - *Prince*

A couple of parents expressed they were uncomfortable with the sexual content in the show and wished the series could include storylines on abstinence.

> I’m a Christian man, so having sexual relations outside of marriage, that I do not teach my children. […] Normalising this thing makes adults more uncomfortable and will create a negative impact on the world we live in. It makes us uncomfortable because we grew up in a time where abstinence is key. –*Parent*

Many parents said they wanted to use MTV Shuga to educate their children about protecting themselves against HIV and getting tested. This is perhaps in contrast to the discussions young people want to have, which would be more open and nuanced, where children and parents can learn from each other.

#### Digital inaccessibility made it difficult for some to watch and discuss

Contextual factors made it difficult for some young people to be fully immersed in the series, most notably lack of a reliable internet connection or WIFI and the cost of data. Unreliable networks and limited data resulted in missed episodes or scenes. Kungawo explained people ‘have data problems or connection problems. Some won’t be patient with the connection and will end up not watching the show.’ Lack of access to TVs and Internet data, either because of unreliable internet or lack of money to pay for data, was a barrier to discussions, as friends may not have the ability to access the show. ‘I never spoke with anyone because there are few houses with TV so I don’t want to look like I’m showing off with the fact that I have a TV.’ - *Junior*

## Discussion

Young people in Mthatha tuned into Shuga because it was a trusted source of information about sexual health. MTV Shuga’s tolerant and empathetic approach to HIV education differed from the “force-fed” information they learned in schools and their parents’ lectures. MTV Shuga allowed space for ambivalence and uncertainty as young people wrestled with topics about sexual health before or as they experienced them personally. Through MTV Shuga storylines, young people experienced a learning journey as they watched realistic characters in relatable contexts wrestle with sexual health decisions while balancing outside influences like studies, financial struggles and relationships. The characters’ situations were not easily resolved, which inspired participants to reflect on and debate how they personally would approach the situations, actively engaging them in the education process. MTV Shuga facilitated conversations about sexual health in homes, schools and university among people who, participants reported, do not usually talk about sexual health. Discussions created valuable sexual health support systems, especially among peers. Watching Shuga with others allowed young people to assess if it was safe to disclose personal information like HIV status and sexual orientation to the person(s) they watched with, based on their reaction to the show.

Participants’ motivation for engaging with MTV Shuga can help us understand what some young people want from HIV education. Using dynamic storytelling in HIV education resonates with young people because stories can reflect that sexual health decisions are complex and intersect with other life choices. Storytelling can guide young people through sexual health situations, educating them about HIV resources and services plus strategies to reduce the risk of HIV. With these resources and knowledge, young people feel empowered to form their own opinions, preparing them for their future sexual relationships. Young people desire autonomy to reflect on HIV choices in a space tolerant of indecision and diverse preferences, as many young people are making decisions about their sexual health for the first time.

Another motivation for engaging with MTV Shuga was the open-ended discussions it inspired, leading to shared decision-making and safe disclosure. Broadcasting and streaming in homes and university residences inspired conversations about sex and HIV that likely would not have occurred otherwise. These findings showed the value of disseminating HIV education beyond classrooms and clinics and more broadly into communities. Having mass media resources on hand allows young people to educate and influence their partners and other meaningful relationships. Mass media can also target out-of-school stakeholders, who benefit from HIV education. However, young people may resist engaging in sexual health mass media interventions in homes. Though young people who discuss sexual health with parents have better outcomes (Biddlecom, Awusabo-Asare, and Bankole 2009; DiClemente et al. 2001; Hutchinson et al. 2003; Flores and Barroso 2017; Coetzee et al. 2014; Martino et al. 2008) many avoid these discussions to prevent judgemental and awkward conversations. Parents struggle to initiate talks and could benefit from receiving updated sexual health information. But, young people worry that making interventions ‘parent appropriate’ would result in less candid and relatable content. Separate media resources tailored to parents could help parents be empathetic and educated advisors to their children.

Despite being digitally connected, participants still reported limited internet and TV access as barriers to watching MTV Shuga and referring it to friends. Digital education and TV interventions likely leave out some of the most vulnerable youth (Adarkwah 2020; Walters 2020). A 2020 study from the Eastern Cape of South Africa reports a positive correlation between media use and HIV knowledge(Shamu et al. 2020). Integrating mass media into schools can make access to media interventions more equitable, as school-based sex education is a cost-effective, practical approach to reaching large numbers of young people from diverse social backgrounds(UNESCO June 2009). Media resources can provide engaging content with clear, tolerant messages about sexual health and assist teachers who do not feel comfortable or lack the training to deliver sexual health education alone(Mturi and Bechuke 2019; Ahmed et al. 2009). The MTV Shuga campaign includes a school-based peer education programme that uses video clips to teach HIV education and inspire conversations. Outside of classrooms, making internet and electricity more affordable and reliable while investing in offline interventions like radio series and comic books will make engaging story-based HIV education more accessible. TV-based interventions should be broadcast when youth are home, available, and control the TV.

### Limitations

Participants in this study were recruited through an online survey, likely attracting digitally connected youth. Many of our participants said they had watched the series on YouTube, which meant they were likely to have reliable internet access, though they also discussed the challenges with internet connectivity. We also did not include those exposed to only the graphic novel, perhaps leaving out less digitally connected youth. Though young people regularly use the internet and digital devices in the Eastern Cape (*South Africa Demographic and Health Survey* 2016:), internet data is expensive.

Uncapped Wi-Fi is difficult to access unless attending higher education, which could sway our sample to a higher income and more educated population. Our participants had sought out MTV Shuga and may have different HIV promotion and education preferences than those who did not engage with MTV Shuga. Those who opted-in to further research on MTV Shuga may also represent the most enthusiastic viewers, and we may have missed the perspectives of those with less interest in the show. Finally, many young people were resistant to referring their parents to the study. Likely, those who did make referrals were already relatively comfortable discussing sexual health with their parents. Hence, this study’s parents may represent a more open group and likely to discuss sexual health than other parents.

### Reflection on remote methodology

Remote data collection offered challenges and benefits in the research process. Phone interviews were a successful, flexible approach to in-depth interviews allowing young people and parents to be interviewed at their convenience, between class and work schedules. However, nonverbal communications are missed over the phone. WhatsApp interviews were flexible and could be conducted asynchronously; however, they did not produce rich data as probing was difficult over text and voice notes. Focus groups conducted over Zoom allowed groups to meet without travelling, but, younger participants struggled with connecting to Zoom because of internet, audio and battery issues, disrupting the discussions. Data collectors asked participants if they were in a safe private place to ensure genuine responses, but they were unable to ensure this was always the case.

Remote methods affected the dynamics between participants and data collectors. Data collectors were selected based on their age to ensure that young people felt comfortable giving candid responses to interview questions. During telephone interviews, where young participants could not see the interviewer, participants were formal and overwhelmingly positive of MTV Shuga and HIV prevention technologies. In focus groups where participants could see the data collectors, conversations were more relaxed and candid, with young people feeling comfortable sharing honest opinions.

## Conclusion

Young viewers engage with MTV Shuga because it guides them through complex sexual health scenarios using stories of relatable young people. Sexual health stories inspire young viewers to reflect on, discuss, and debate sexual health. Young people tuned in to MTV Shuga because of its tolerant tone, which allowed space for uncertainty and diverse opinions about HIV and other sexual health topics. Reflection and conversations plus greater awareness about the sexual health services available can prepare young people for their future sexual lives. Mass media interventions like MTV Shuga can disseminate resources and knowledge into communities’ and help to initiate conversations about HIV among family, friends and partners. Teachers who deliver sexual health education can also utilise mass media for engaging HIV education content. Scaling up access to offline story-based media while increasing young people’s accessibility to the internet and television is essential for making engaging HIV education more accessible.

## Data Availability

All data produced in the present study are available upon reasonable request to the authors

## Acknowledgements

The authors wish to thank all the young people who contributed their time to this study. We especially would like to thank Luntu Fica, Bawelile Madlala, Samkele Khuma, and Lunga Mkrezo for their hard work and valuable ideas which strengthen this research. We are grateful to Dominique O’Donnell, Antonio Duran-Aparicio and Sithembele Ntenteni for all the support to make this project possible. We acknowledge the dedication of MTV Shuga peer educators and coordinators (Yvonne Diogo, Lesedi Thwala) in South Africa and appreciate the trust of the MTV Staying Alive Foundation, especially Alistair Chase, Georgia Arnold and Sara Piot.

